# White matter lesions as a prognostic marker of recurrence in cryptogenic stroke with high-risk patent foramen ovale

**DOI:** 10.1101/2023.09.03.23295000

**Authors:** Shunichi Niiyama, Yuji Ueno, Naohide Kurita, Sho Nakajima, Chikage Kijima, Kenichiro Hira, Nobukazu Miyamoto, Masao Watanabe, Kazuo Yamashiro, Takao Urabe, Nobutaka Hattori

## Abstract

****Introduction**:** Atrial septal aneurysms increase the risk of stroke recurrence in cryptogenic stroke patients with a patent foramen ovale. Factors related to stroke recurrence according to the patent foramen ovale risk stratification have not been fully evaluated.

****Methods**:** Data from a multicenter observational registry of ischemic stroke patients undergoing transesophageal echocardiography, with a comprehensive database including clinical characteristics and long-term prognosis, were used for this study. Patients were classified into three groups: high-risk patent foramen ovale group, large shunt patent foramen ovale (≥20 microbubbles) or patent foramen ovale with atrial septal aneurysm; right-to-left shunt group, right-to-left shunt including patent foramen ovale with <20 microbubbles or without atrial septal aneurysm; and negative right-to-left shunt group. Among stroke subtype, factors related to stroke recurrence were investigated according to the risk stratification of patent foramen ovale in cryptogenic stroke.

****Results**:** In total, 586 patients (185 females; 65.5 ± 13.2 years) were analyzed. In cryptogenic stroke (329 patients) with median follow-up of 4.2 (interquartile range, 1.0– 6.1) years, 55 patients had stroke recurrence. The negative right-to-left shunt, right-to-left shunt, and high-risk patent foramen ovale groups included 179, 90, and 60 patients, in which stroke recurrence occurred in 5.3%, 2.5%, and 4.6% per person-year, respectively. In patients with high-risk patent foramen ovale, the NIH stroke scale score (hazard ratio 1.257, 95% CI 1.034-1.530, *P*=0.022) and periventricular hyperintensity (hazard ratio 3.369, 95% CI 1.103-10.294, *P*=0.035) were predictive markers for stroke recurrence on multivariate Cox Hazards analysis, but no factors were related to stroke recurrence in the right-to-left shunt and negative right-to-left shunt groups.

****Conclusion**:** High-risk patent foramen ovale is an important embolic source in cryptogenic stroke. Periventricular hyperintensity was shown to predict recurrent stroke in patients with a high-risk patent foramen ovale. In such cases, percutaneous patent foramen ovale closure might be effective at <60 years of age.

## Introduction

The foramen ovale, a remnant of the fetal circulation, can still be open in about 25% of the general adult population.^1, 2^ A patent foramen ovale (PFO) has been shown to be a risk factor for ischemic stroke, particularly in young subjects. A recent meta-analysis showed that a PFO was found in 29% of cryptogenic stroke case, two times higher than that in patients with stroke of known cause.^3^ Furthermore, the coexistence of an atrial septal aneurysm increased the risk of stroke recurrence in patients with a PFO.^4^ Meanwhile, a PFO has been shown to increase the risk of recurrent stroke even in elderly subjects.^5^ Transesophageal echocardiography (TEE) is not only the gold standard for detecting a PFO with a degree of right-to-left (RLS) shunt, but also useful for other cardiac anomalies such as atrial septal aneurysms (ASAs).^6^ It has been shown that a high-risk PFO, assessed by TEE as having an ASA and high shunt volume, was a predictor for stroke recurrence, in which PFO closure was useful.^7–10^ So far, predictors of stroke recurrence in cryptogenic stroke patients according to the PFO risk stratification have not been fully studied.

A multicenter registry enrolled patients with brain infarction and transient ischemic attack (TIA) with a comprehensive database including baseline characteristics, laboratory data, radiological findings, therapy, and long-term prognosis, for whom potential embolic etiologies including PFO and ASA were evaluated by TEE. Taking advantage of this registry, the factors related to recurrent stroke in cryptogenic stroke patients with high-risk PFO, with RLS other than high-risk PFO, and without RLS, were investigated.

## Methods

### Study population

The registry was a multicenter, observational registry retrospectively enrolling consecutive patients with ischemic stroke who underwent TEE in Juntendo University Hospital and Juntendo University Urayasu Hospital from February 2012 to December 2017 (COCTAIL-TEE [Clinical Observation, Characteristics, and prognosis for long-term of stroke patients undergoing TransEsophageal Echocardiography]). The inclusion criteria for this registration were as follows: (1) patients with brain infarction and TIA, (2) stroke patients underwent TEE. The patients’ medical history and diagnostic modalities including magnetic resonance imaging (MRI), 12-lead electrocardiography, blood examinations, and TEE performed during admission were evaluated. Treatment with anti-thrombotic agents and long-term outcomes such as stroke recurrence after hospital discharge were retrospectively analyzed. The institutional review boards of both participating hospitals approved the study protocol. Clinical information obtained from medical records was used; thus, the need to obtain written, informed consent from each patient was waived in this retrospective study. This study was conducted in accordance with the Declaration of Helsinki.

### Atherosclerotic risk factors

Hypertension was defined as systolic blood pressure >140 mmHg or diastolic blood pressure >90 mmHg after the 14th admission day or use of antihypertensive drugs. Diabetes mellitus was defined as hemoglobin A1c levels ≥6.5% on admission or the use of antidiabetic drugs. Dyslipidemia was defined as low-density lipoprotein cholesterol ≥ 140 mg/dl, high-density lipoprotein cholesterol < 40 mg/dl, triglycerides ≥ 150 mg/dl, or the use of lipid-lowering drugs. Ischemic heart disease included myocardial infarction, angina pectoris, and coronary artery stenosis. Chronic kidney disease was defined as an estimated glomerular filtration rate (eGFR) < 60 ml/min/1.73 m^2^.

### MRI sequences

MRI was performed at each institution with 1.5- or 3-T scanners during hospitalization. Sequences included axial diffusion-weighted imaging (DWI), fluid-attenuated inversion recovery imaging (FLAIR), magnetic resonance angiography (MRA), and the GRE T2* sequence. DWI (repetition time (TR) / echo time (TE) =4156-7000/60-120 ms) was used to assess the size and distribution of the index stroke lesions. FLAIR (TR/TE =6879-10000/105-120 ms) was used to evaluate the degree of deep and subcortical white matter hyperintensity (DSWMH) and periventricular hyperintensity (PVH) in accordance with Fazekas grades 0-3.^11^ MRA (TR/TE = 19-30/2.8-6.9 ms) was used to detect intracranial stenosis >50% not relevant to the infarction area. Examinations were performed by two or three experienced sonographers in each institution.

### TEE study

TEE was performed as explained in our previous work.^12^ An ASA was defined as a ≥10-mm excursion into either the left or right atrium, or a sum of total excursion into the left or right atrium of ≥15 mm. The right-to-left shunt (RLS) was assessed by injecting agitated saline and having patients perform the Valsalva maneuver.

Subsequently, the number of microbubbles with and without contrast agents was compared, and the number transiting from the right to the left atrium was counted. PFO was diagnosed when microbubbles were visualized in the left atrium ≤3 cardiac cycles after the Valsalva maneuver, whereas a pulmonary arteriovenous fistula was diagnosed when microbubbles were visualized in the left atrium >3 cardiac cycles after the Valsalva maneuver or when microbubbles were visualized without the Valsalva maneuver.

### Classification of patients based on the presence of RLS

Based on the presence or absence of RLS, patients were classified into three groups: negative RLS group, lacking RLS; RLS group, RLS including a PFO or pulmonary arteriovenous fistula with a small-to-moderate shunt (<20 microbubbles) or without ASA; and high-risk PFO group, having a large-shunt PFO (≥20 microbubbles) or concurrent ASA.

### Data collection for analyses and outcome measures

Baseline clinical information and radiological and echocardiographic data during hospitalization were collected through hospital charts or database reviews. During the study period from May 2021 to December 2021, such data were retrospectively obtained. The primary outcome of stroke recurrence, for any type of stroke occurrence such as brain infarction, transient ischemic attack, intracranial hemorrhage, and subarachnoid hemorrhage, was also extracted from hospital charts or database reviews. In enrolled patients, ischemic stroke subtype was determined according to the Trial of Org 10172 in Acute Stroke Treatment (TOAST) criteria.^13^ Clinical characteristics were compared between cryptogenic stroke and other stroke subtype, and factors related to stroke recurrence were investigated in the negative RLS, RLS, and high-risk PFO groups, in cryptogenic stroke.

### Statistical analysis

Data are presented as number (%), mean ± standard deviation, and median (interquartile range) values. Patients’ characteristics and baseline data were compared between the two groups. Continuous variables were compared using the Mann-Whitney test, and nominal variables were analyzed by the chi-squared test. The incidence of recurrent stroke was estimated per 100 person-years of observation by negative RLS, RLS, and high-risk PFO in cryptogenic stroke. The Cox proportional hazards model was used to explore factors associated with recurrent stroke. After univariate analysis of all clinically relevant covariates, those with a *P*<0.2 were included in the multivariable Cox model. The Kaplan-Meier method and log-rank test were used to estimate cumulative event rates of recurrent vascular events. Missing data were handled with pairwise deletion. Significance (*P* value) was set at <0.05. Statistical analyses were conducted using IBM SPSS Statistics version 27 (IBM, Armonk, NY, USA).

## Results

A total of 598 patients with ischemic stroke and TIA were enrolled in the COCTAIL-TEE registry from two hospitals in Japan. Of these, seven patients whose follow-up data were unavailable and five patients who were not evaluated for the presence of RLS during TEE examinations were excluded. Thus, the data of 586 patients (185 females; 65.5 ± 13.2 years) with median follow-up of 4.2 (interquartile range, 1.0–6.3) years were analyzed (Fig. 1).

**Figure 1.**
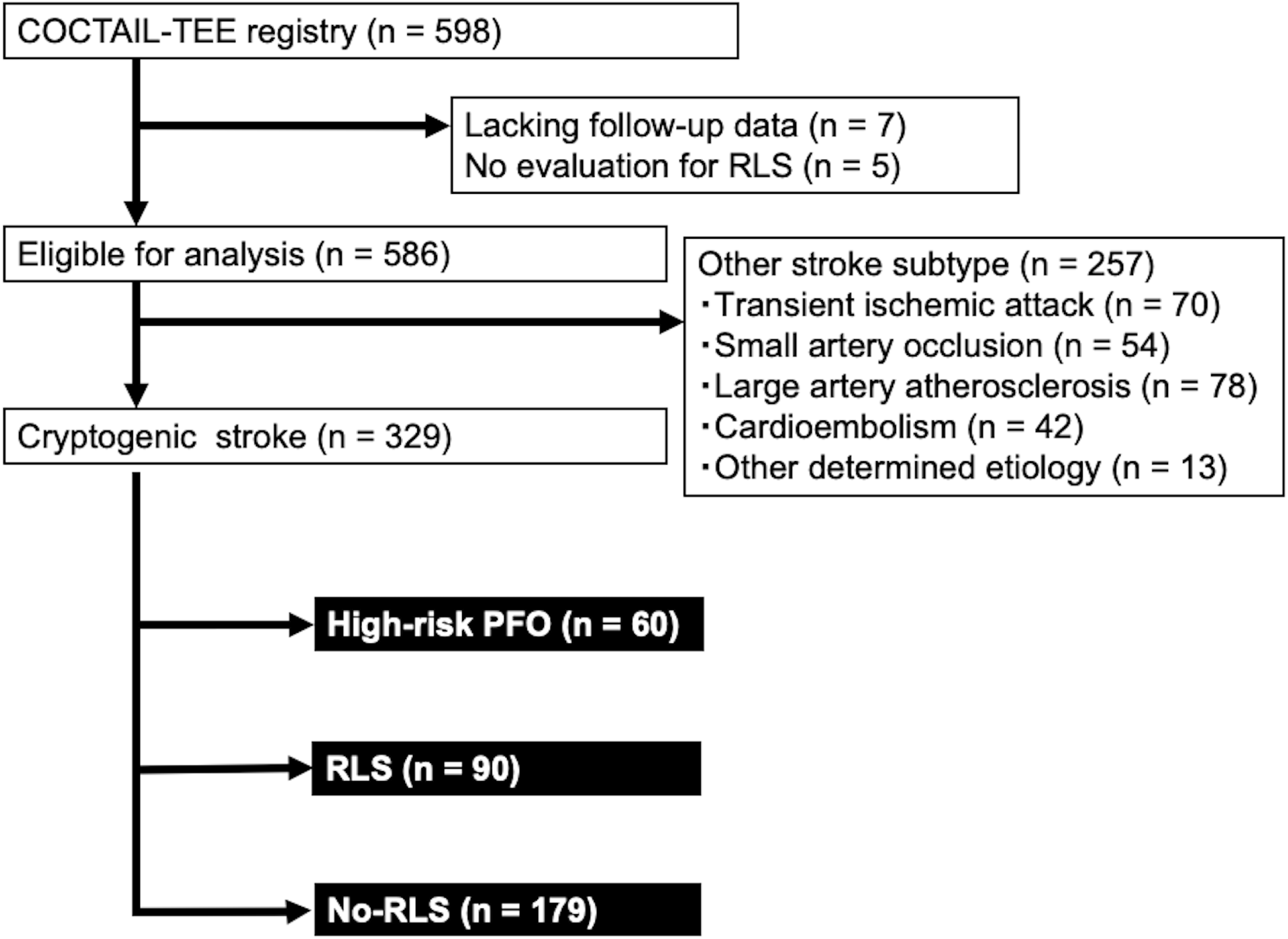
COCTAIL-TEE = Clinical Observation, Characteristics, and prognosis for long-term of stroke patients undergoing TransEsophageal Echocardiography, RLS = right-to-left shunt, PFO = patent foramen ovale.

### Clinical characteristics of patients with cryptogenic stroke and other stroke subtype

Overall, 329 patients were categorized as having cryptogenic stroke, and 257 patients had other stroke subtypes, consisting of 70 patients with transient ischemic attack, 54 patients with small artery occlusion, 78 patients with large artery atherosclerosis, 42 with cardioembolism, and 13 with other determined etiology including branch atheromatous disease (Fig. 1). Table 1 summarizes the clinical characteristics of patients with cryptogenic stroke compared with other stroke subtypes. Age, frequency of female sex, atherosclerotic risk factors, NIHSS score, degrees of PVH and DSWMH, and levels of glucose and LDL-C were not significantly different between cryptogenic stroke and other stroke subtypes. D-dimer levels were lower in cryptogenic stroke than in other stroke subtypes (2.0 ± 4.2 vs. 2.3 ± 12.8 μg/mL, *P*=0.048) (Table 1).

**Table 1.**
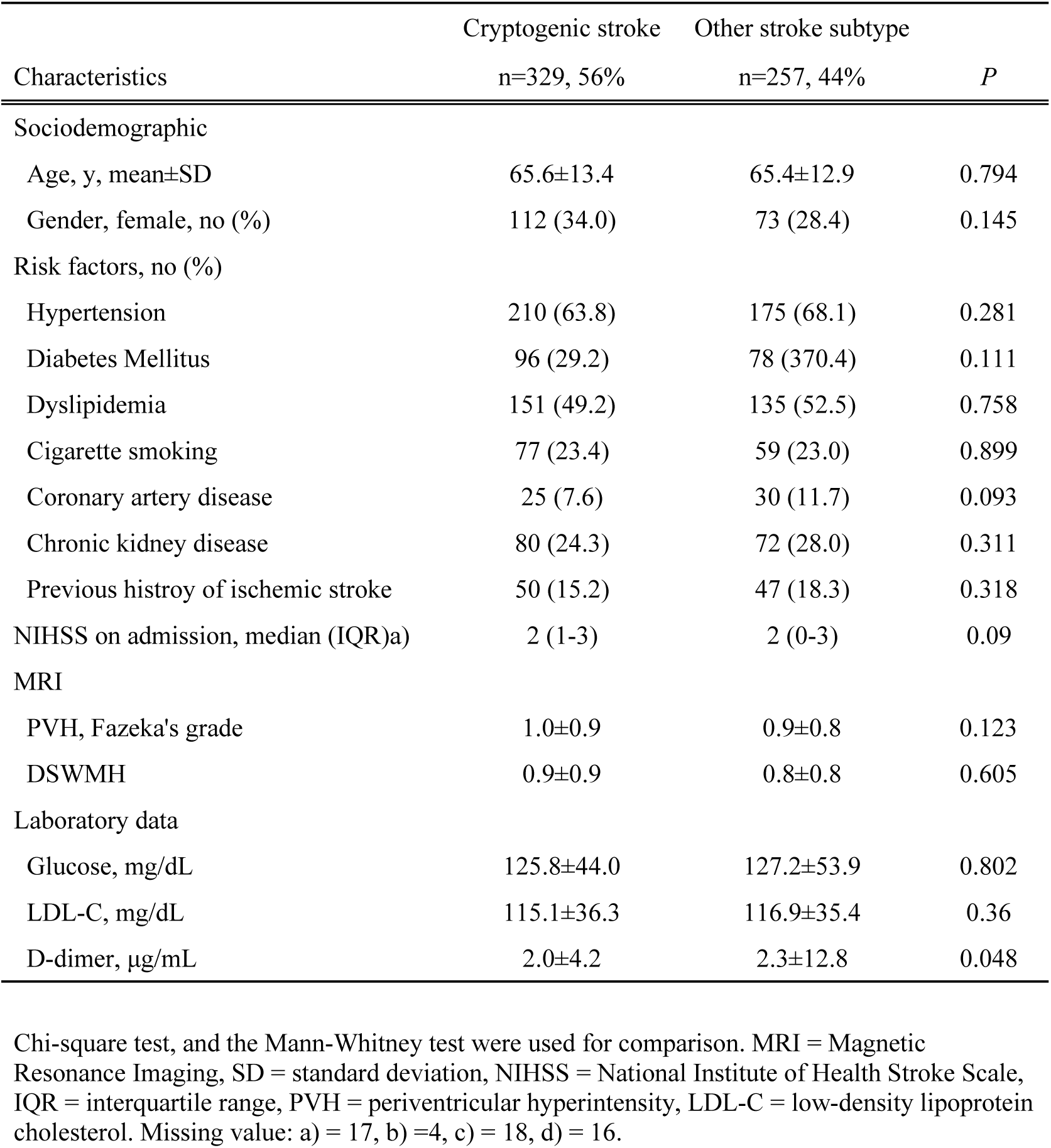
Baseline characteristics, MRI findings, and laboratory data of the cryptogenic stroke and other stroke subtype groups.

### Clinical characteristics of cryptogenic stroke patients with stroke recurrence in the negative RLS, RLS, and high-risk PFO groups

In cryptogenic stroke, the median follow-up was 4.2 (interquartile range, 1.0–6.1) years, and 55 patients had stroke recurrence. The negative RLS, RLS, and high-risk PFO groups included 179, 90, and 60 patients, respectively (Fig. 1). Table 2 shows that there were 35 patients with recurrent stroke in the negative RLS group. The frequency of a previous history of ischemic stroke was relatively higher in patients with stroke recurrence (25.7% vs. 13.9%, *P*=0.089), whereas the frequency of large infarct >3.0 cm in diameter was relatively lower in patients with stroke recurrence, compared to patients without stroke recurrence (14.3% vs. 27.8%, *P*=0.098). In the RLS group, 9 patients had recurrent stroke, and the degrees of PVH and DSWMH were significantly higher in patients with stroke recurrence than in patients without stroke recurrence (1.8±0.8 vs. 0.8±0.9, *P*=0.002; 1.7±0.9 vs. 0.7±0.8, *P*=0.001). In the high-risk PFO group, 11 patients had stroke recurrence, and the NIHSS score on admission (2 [1.5-4.5] vs. 1 [0-3], *P*=0.053) was relatively higher than in patients without stroke recurrence. Degrees of PVH and DSWMH were significantly higher in patients with stroke recurrence than in patients without stroke recurrence (1.7±1.0 vs. 0.8±1.0, *P*=0.003; 1.3±1.0 vs. 0.7±0.8, *P*=0.041).

**Table 2.**
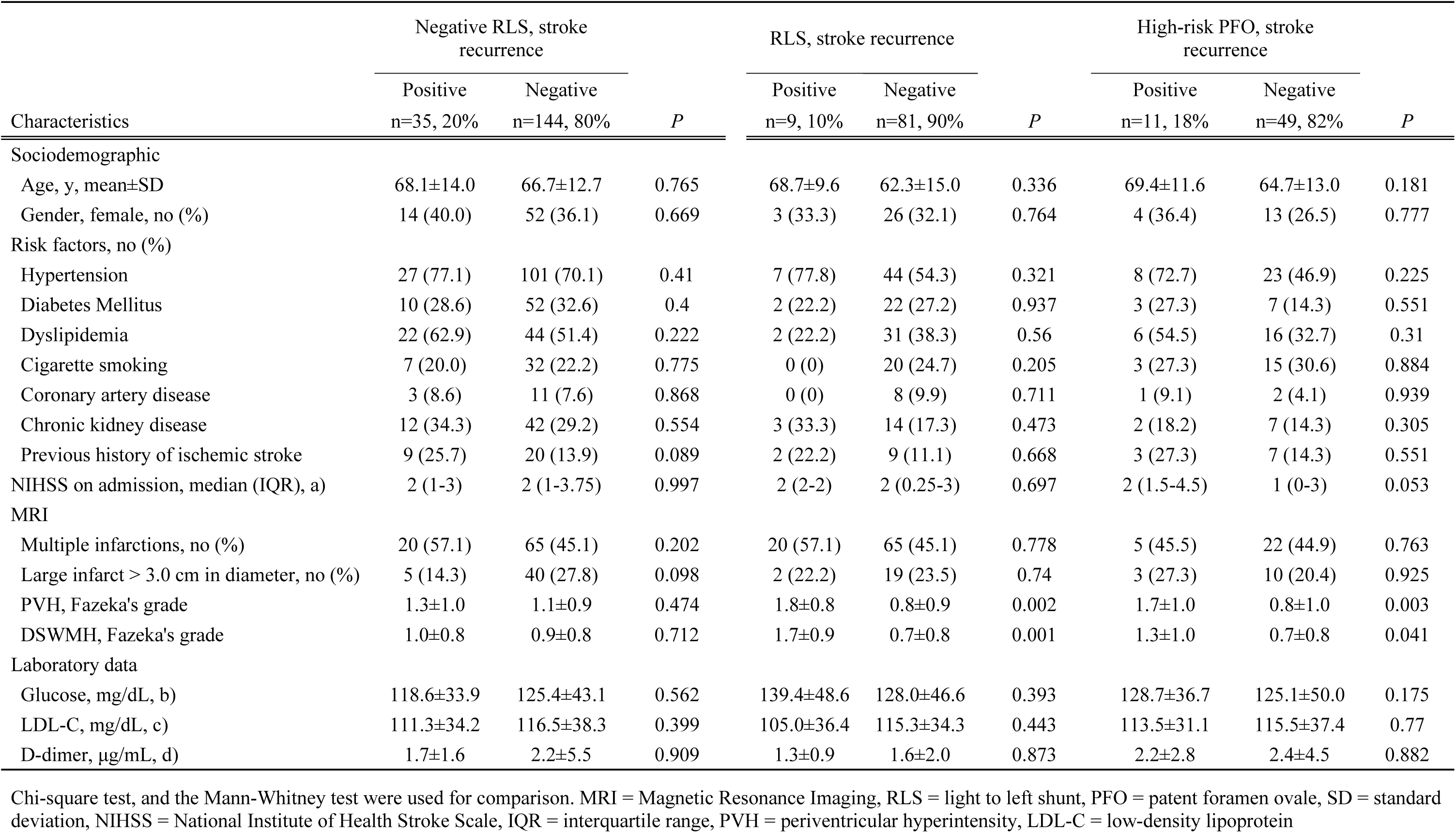
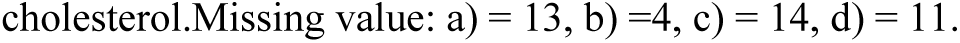
Baseline characteristics, MRI findings, and laboratory data according to the presence of stroke recurrence in cryptogenic stoke with negative RLS, RLS, and high-risk PFO.

| Characteristics | Negative RLS, stroke recurrence |  |  | RLS, stroke recurrence |  |  | High-risk PFO, stroke recurrence |  |  |
| --- | --- | --- | --- | --- | --- | --- | --- | --- | --- |
|  | Positive<br>n=35, 20% | Negative<br>n=144, 80% | *P* | Positive<br>n=9, 10% | Negative<br>n=81, 90% | *P* | Positive<br>n=11, 18% | Negative<br>n=49, 82% | *P* |
| **Sociodemographic** |  |  |  |  |  |  |  |  |  |
| Age, y, mean±SD | 68.1±14.0 | 66.7±12.7 | 0.765 | 68.7±9.6 | 62.3±15.0 | 0.336 | 69.4±11.6 | 64.7±13.0 | 0.181 |
| Gender, female, no (%) | 14 (40.0) | 52 (36.1) | 0.669 | 3 (33.3) | 26 (32.1) | 0.764 | 4 (36.4) | 13 (26.5) | 0.777 |
| **Risk factors, no (%)** |  |  |  |  |  |  |  |  |  |
| Hypertension | 27 (77.1) | 101 (70.1) | 0.41 | 7 (77.8) | 44 (54.3) | 0.321 | 8 (72.7) | 23 (46.9) | 0.225 |
| Diabetes Mellitus | 10 (28.6) | 52 (32.6) | 0.4 | 2 (22.2) | 22 (27.2) | 0.937 | 3 (27.3) | 7 (14.3) | 0.551 |
| Dyslipidemia | 22 (62.9) | 44 (51.4) | 0.222 | 2 (22.2) | 31 (38.3) | 0.56 | 6 (54.5) | 16 (32.7) | 0.31 |
| Cigarette smoking | 7 (20.0) | 32 (22.2) | 0.775 | 0 (0) | 20 (24.7) | 0.205 | 3 (27.3) | 15 (30.6) | 0.884 |
| Coronary artery disease | 3 (8.6) | 11 (7.6) | 0.868 | 0 (0) | 8 (9.9) | 0.711 | 1 (9.1) | 2 (4.1) | 0.939 |
| Chronic kidney disease | 12 (34.3) | 42 (29.2) | 0.554 | 3 (33.3) | 14 (17.3) | 0.473 | 2 (18.2) | 7 (14.3) | 0.305 |
| Previous history of ischemic stroke | 9 (25.7) | 20 (13.9) | 0.089 | 2 (22.2) | 9 (11.1) | 0.668 | 3 (27.3) | 7 (14.3) | 0.551 |
| NIHSS on admission, median (IQR), a) | 2 (1-3) | 2 (1-3.75) | 0.997 | 2 (2-2) | 2 (0.25-3) | 0.697 | 2 (1.5-4.5) | 1 (0-3) | 0.053 |
| **MRI** |  |  |  |  |  |  |  |  |  |
| Multiple infarctions, no (%) | 20 (57.1) | 65 (45.1) | 0.202 | 20 (57.1) | 65 (45.1) | 0.778 | 5 (45.5) | 22 (44.9) | 0.763 |
| Large infarct > 3.0 cm in diameter, no (%) | 5 (14.3) | 40 (27.8) | 0.098 | 2 (22.2) | 19 (23.5) | 0.74 | 3 (27.3) | 10 (20.4) | 0.925 |
| PVH, Fazeka's grade | 1.3±1.0 | 1.1±0.9 | 0.474 | 1.8±0.8 | 0.8±0.9 | 0.002 | 1.7±1.0 | 0.8±1.0 | 0.003 |
| DSWMH, Fazeka's grade | 1.0±0.8 | 0.9±0.8 | 0.712 | 1.7±0.9 | 0.7±0.8 | 0.001 | 1.3±1.0 | 0.7±0.8 | 0.041 |
| **Laboratory data** |  |  |  |  |  |  |  |  |  |
| Glucose, mg/dL, b) | 118.6±33.9 | 125.4±43.1 | 0.562 | 139.4±48.6 | 128.0±46.6 | 0.393 | 128.7±36.7 | 125.1±50.0 | 0.175 |
| LDL-C, mg/dL, c) | 111.3±34.2 | 116.5±38.3 | 0.399 | 105.0±36.4 | 115.3±34.3 | 0.443 | 113.5±31.1 | 115.5±37.4 | 0.77 |
| D-dimer, µg/mL, d) | 1.7±1.6 | 2.2±5.5 | 0.909 | 1.3±0.9 | 1.6±2.0 | 0.873 | 2.2±2.8 | 2.4±4.5 | 0.882 |

Chi-square test, and the Mann-Whitney test were used for comparison. MRI = Magnetic Resonance Imaging, RLS = light to left shunt, PFO = patent foramen ovale, SD = standard deviation, NIHSS = National Institute of Health Stroke Scale, IQR = interquartile range, PVH = periventricular hyperintensity, LDL-C = low-density lipoprotein
cholesterol.Missing value: a) = 13, b) =4, c) = 14, d) = 11.

### Factors associated with stroke recurrence according to the presence of RLS

The incidence of stroke recurrence in the three groups was 5.3%, 2.5%, and 4.6% per person-year, respectively. In the negative RLS group, dyslipidemia, previous history of ischemic stroke, multiple infarctions, and large infarct > 3.0 cm reached *P*<0.2 on univariate Cox hazards analysis and were entered into the multivariate Cox hazards analysis. However, no factors were found to predict stroke recurrence (Table 3). In the RLS group, the univariate Cox hazards analysis showed that PVH and DSWMH significantly predicted stroke recurrence (hazard ratio [HR] 2.319, 95% CI 1.257-4.277, *P*=0.007; HR 2.506, 95% CI 1.349-4.654, *P*=0.004). However, multivariate analysis found no factors predicting stroke recurrence (Table 4). In the high-risk PFO group, NIHSS score on admission (HR 1.346, 95% CI 1.147-1.579, *P*<0.001), PVH (HR 2.520, 95% CI 1.417-4.484, *P*=0.002), and DSWMH (HR 2.547, 95% CI 1.296-5.008, *P*=0.007) showed significant differences and were entered into the multivariate Cox hazards model. The NIHSS score on admission (HR 1.257, 95% CI 1.034-1.530, *P*=0.022) and PVH (HR 3.369, 95% CI 1.103-10.294, *P*=0.035) were found to be predictive markers for stroke recurrence (Table 5). The Kaplan-Meier survival curves showed that cumulative event-free rates were significantly lower in patients with a higher degree of PVH in patients with RLS (log-rank test, *P*=0.029) and with high-risk PFO (log-rank test, *P*=0.004) (Fig. 2).

**Figure 2.**
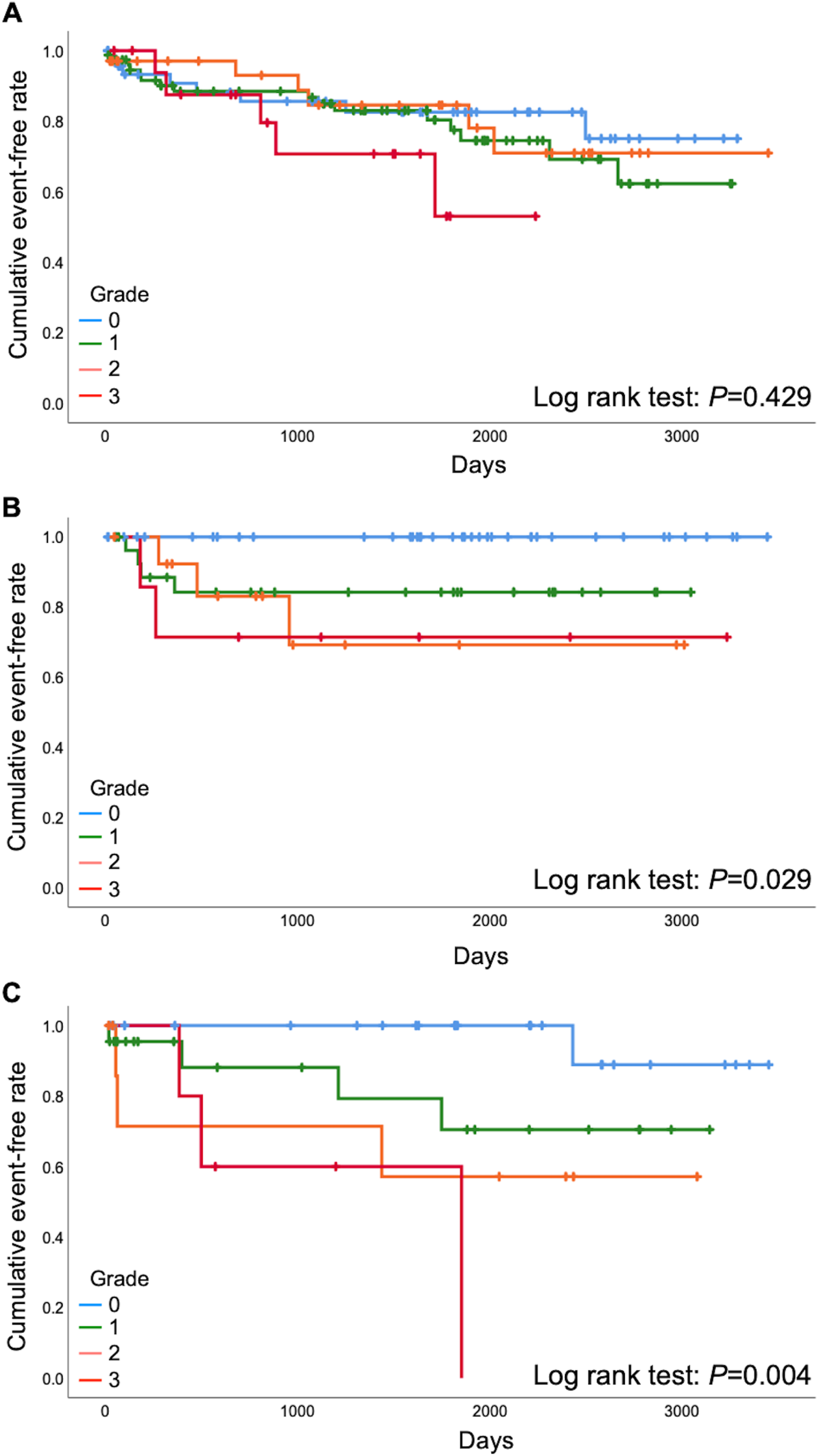
Kaplan-Meier curves of freedom from stroke recurrence during follow-up. The X axis indicates time in days since inclusion in the study. The Y axis indicates the proportion of patients surviving free of stroke recurrence. Cumulative event-free rates are compared among degrees of periventricular hyperintensity in the negative RLS (**A**), RLS (**B**), and high-risk PFO (**C**) groups. RLS = right-to-left shunt, PFO = patent foramen ovale.

**Table 3.** Univariate and multivariate analyses of baseline characteristics, MRI findings, and laboratory data predicting stroke recurrence in cryptogenic stroke without RLS.

| Characteristics | Univariate analysis |  |  | Multivariate analysis |  |  |
| --- | --- | --- | --- | --- | --- | --- |
|  | HR | 95% CI | *P* | HR | 95% CI | *P* |
| Sociodemographic |  |  |  |  |  |  |
| Age | 1.011 | 0.984-1.039 | 0.417 |  |  |  |
| Gender, female | 1.162 | 0.591-2.288 | 0.663 |  |  |  |
| Risk factors |  |  |  |  |  |  |
| Hypertension | 1.083 | 0.491-2.390 | 0.843 |  |  |  |
| Diabetes Mellitus | 0.727 | 0.349-1.513 | 0.394 |  |  |  |
| Dyslipidemia | 1.606 | 0.808-3.191 | 0.176 | 1.694 | 0.847-3.388 | 0.136 |
| Cigarette smoking | 0.835 | 0.364-1.916 | 0.67 |  |  |  |
| Coronary artery disease | 0.937 | 0.286-3.064 | 0.914 |  |  |  |
| Chronic kidney disease | 1.289 | 0.640-2.596 | 0.477 |  |  |  |
| Previous history of ischemic stroke | 1.925 | 0.899-4.122 | 0.092 | 1.831 | 0.852-3.935 | 0.121 |
| NIHSS on admission, a) | 0.932 | 0.809-1.073 | 0.326 |  |  |  |
| MRI |  |  |  |  |  |  |
| Multiple infarctions | 1.754 | 0.894-3.442 | 0.102 | 1.723 | 0.874-3.396 | 0.116 |
| Large infarct > 3.0 cm in diameter | 0.493 | 0.191-1.272 | 0.143 | 0.477 | 0.184-1.240 | 0.129 |
| PVH | 1.225 | 0.855-1.756 | 0.27 |  |  |  |
| DSWMH | 1.138 | 0.765-1.692 | 0.524 |  |  |  |
| Laboratory data |  |  |  |  |  |  |
| Glucose, b) | 0.994 | 0.985-1.004 | 0.229 |  |  |  |
| LDL-C, c) | 0.997 | 0.987-1.007 | 0.542 |  |  |  |
| D-dimer, d) | 1.023 | 0.925-1.131 | 0.661 |  |  |  |
| Treatment |  |  |  |  |  |  |
| Antiplatelet agent | 1.463 | 0.516-4.146 | 0.474 |  |  |  |
| Anticoagulant therapy | 0.691 | 0.244-1.957 | 0.486 |  |  |  |

MRI = Magnetic Resonance Imaging, RLS = light to left shunt, HR = hazard ratio, CI = confidence interval NIHSS = National Institute of Health Stroke Scale, PVH = periventricular hyperintensity, LDL-C = low-density lipoprotein cholesterol. Missing value: a) = 13, b) = 4, c) = 14, d) = 11.

**Table 4.**
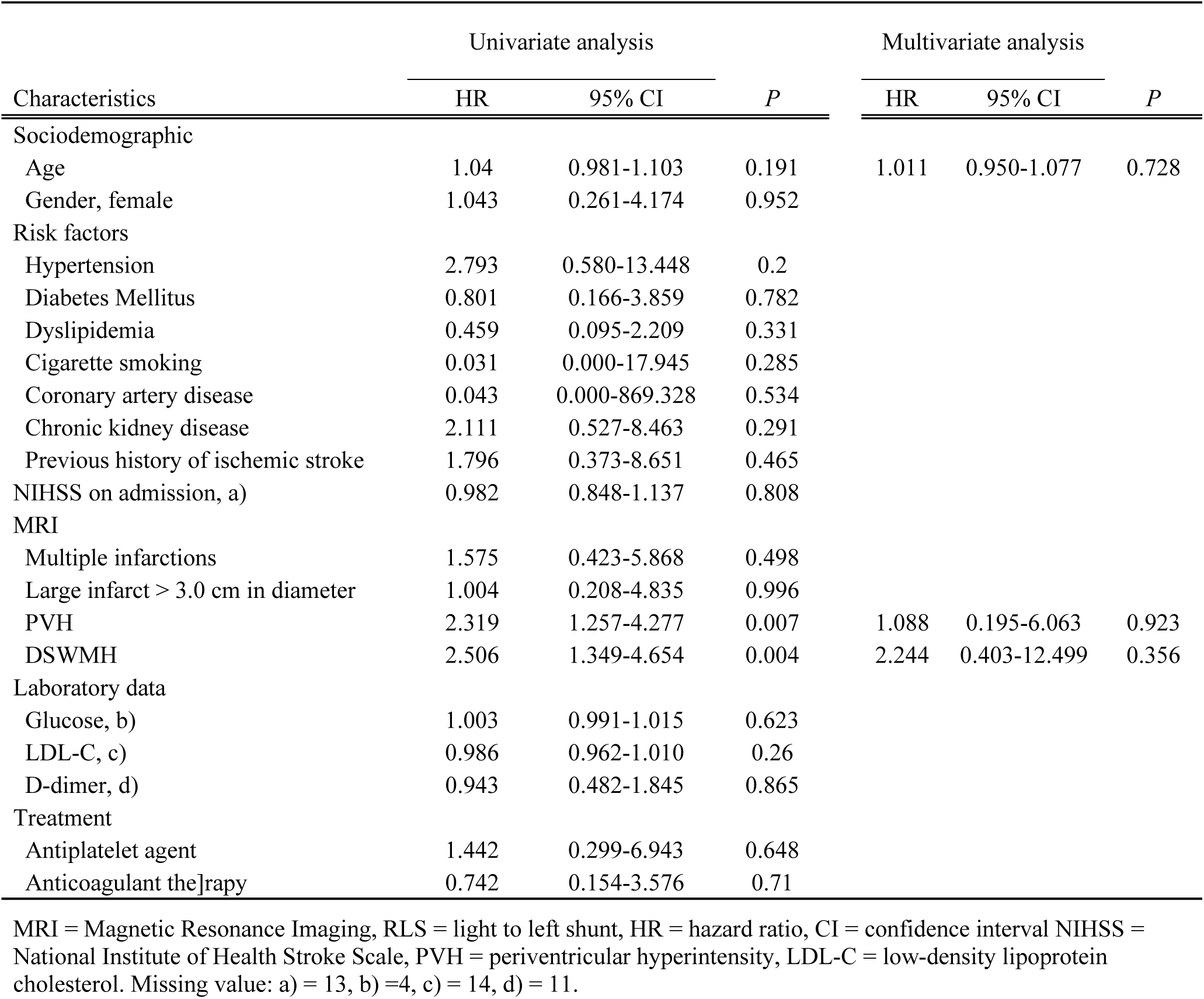
Univariate and multivariate analyses of baseline characteristics, MRI findings, and laboratory data predicting stroke recurrence in cryptogenic stroke with RLS.

| Characteristics | Univariate analysis |  |  | Multivariate analysis |  |  |
| --- | --- | --- | --- | --- | --- | --- |
|  | HR | 95% CI | P | HR | 95% CI | P |
| Sociodemographic |  |  |  |  |  |  |
| Age | 1.04 | 0.981-1.103 | 0.191 | 1.011 | 0.950-1.077 | 0.728 |
| Gender, female | 1.043 | 0.261-4.174 | 0.952 |  |  |  |
| Risk factors |  |  |  |  |  |  |
| Hypertension | 2.793 | 0.580-13.448 | 0.2 |  |  |  |
| Diabetes Mellitus | 0.801 | 0.166-3.859 | 0.782 |  |  |  |
| Dyslipidemia | 0.459 | 0.095-2.209 | 0.331 |  |  |  |
| Cigarette smoking | 0.031 | 0.000-17.945 | 0.285 |  |  |  |
| Coronary artery disease | 0.043 | 0.000-869.328 | 0.534 |  |  |  |
| Chronic kidney disease | 2.111 | 0.527-8.463 | 0.291 |  |  |  |
| Previous history of ischemic stroke | 1.796 | 0.373-8.651 | 0.465 |  |  |  |
| NIHSS on admission, a) | 0.982 | 0.848-1.137 | 0.808 |  |  |  |
| MRI |  |  |  |  |  |  |
| Multiple infarctions | 1.575 | 0.423-5.868 | 0.498 |  |  |  |
| Large infarct > 3.0 cm in diameter | 1.004 | 0.208-4.835 | 0.996 |  |  |  |
| PVH | 2.319 | 1.257-4.277 | 0.007 | 1.088 | 0.195-6.063 | 0.923 |
| DSWMH | 2.506 | 1.349-4.654 | 0.004 | 2.244 | 0.403-12.499 | 0.356 |
| Laboratory data |  |  |  |  |  |  |
| Glucose, b) | 1.003 | 0.991-1.015 | 0.623 |  |  |  |
| LDL-C, c) | 0.986 | 0.962-1.010 | 0.26 |  |  |  |
| D-dimer, d) | 0.943 | 0.482-1.845 | 0.865 |  |  |  |
| Treatment |  |  |  |  |  |  |
| Antiplatelet agent | 1.442 | 0.299-6.943 | 0.648 |  |  |  |
| Anticoagulant therapy | 0.742 | 0.154-3.576 | 0.71 |  |  |  |

MRI = Magnetic Resonance Imaging, RLS = light to left shunt, HR = hazard ratio, CI = confidence interval NIHSS = National Institute of Health Stroke Scale, PVH = periventricular hyperintensity, LDL-C = low-density lipoprotein cholesterol. Missing value: a) = 13, b) = 4, c) = 14, d) = 11.

**Table 5.**
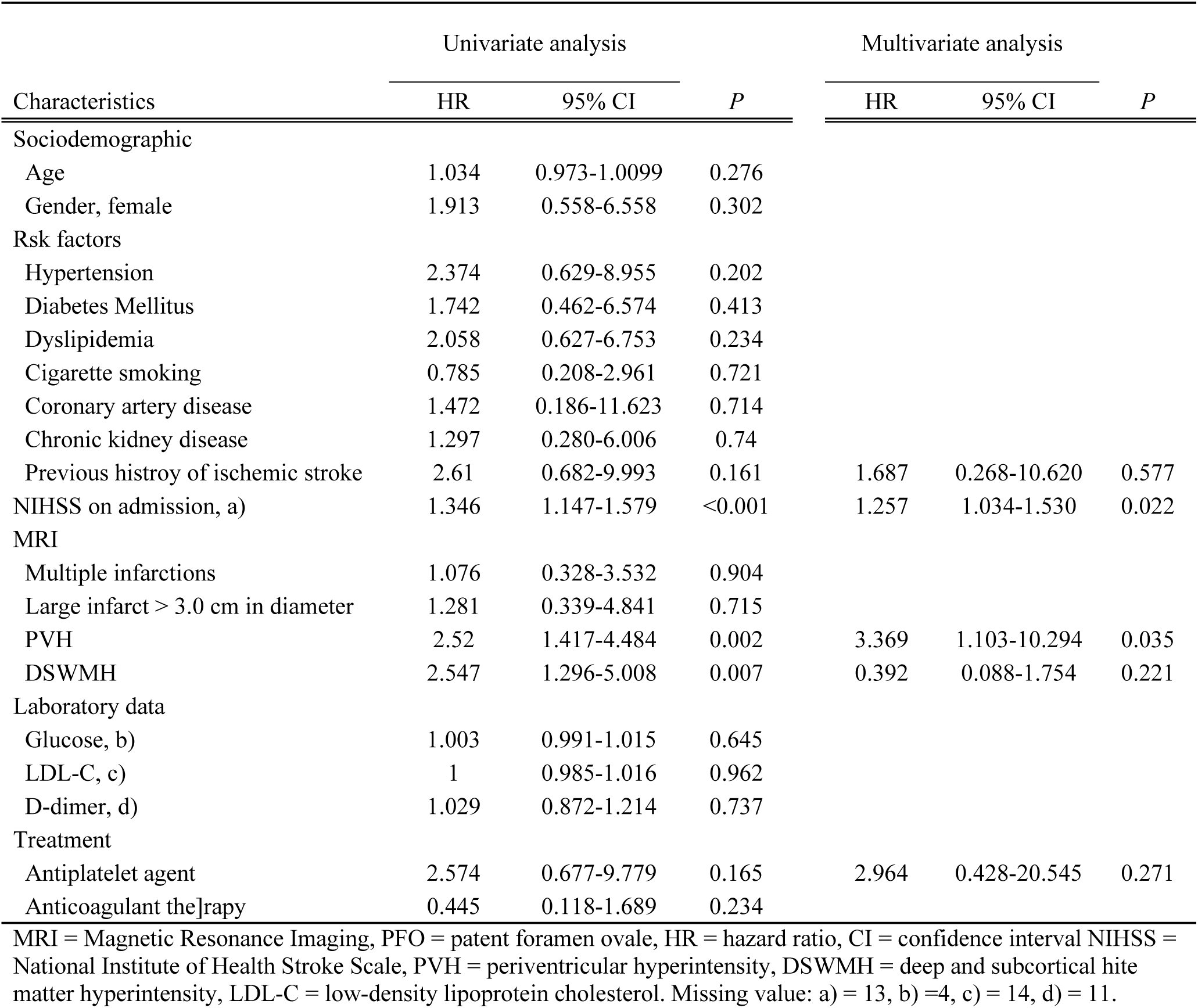
Univariate and multivariate analyses of baseline characteristics, MRI findings, and laboratory data predicting stroke recurrence in cryptogenic stroke with high-risk PFO.

### Treatment for cryptogenic stroke patients according to the presence of RLS

As for secondary prevention of cryptogenic stroke, antiplatelet agents and anticoagulant therapy were given in 86.0% and 15.1%, 72.2% and 27.8%, and 60.0% and 41.7% of the negative RLS, RLS, and high-risk PFO groups, respectively (Fig. 3).

**Figure 3.**
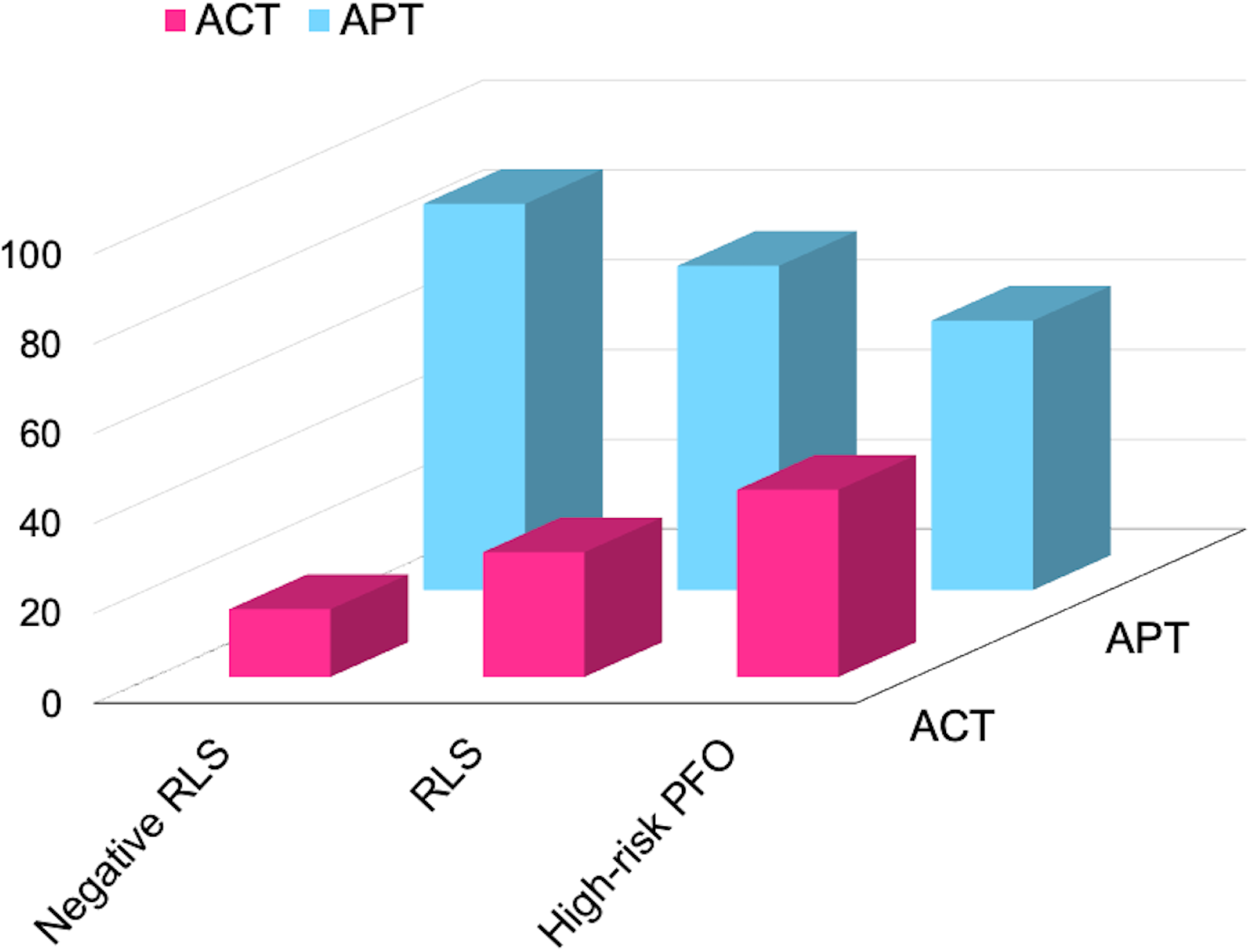
Proportion of antiplatelet and anticoagulant therapies for secondary prevention in the negative RLS, RLS, and high-risk PFO groups. ACT = anticoagulant therapy, APT = antiplatelet therapy, RLS = right-to-left shunt, PFO = patent foramen ovale.

## Discussion

The current study from a multicenter registry of stroke patients undergoing TEE with long-term outcomes provides novel insights into the clinical significance of stroke recurrence in cryptogenic stroke patients according to the presence of RLS. The rate of stroke recurrence was higher in order of negative RLS, high-risk PFO, and RLS. In particular, the presence of white matter lesions was a predictive factor for stroke recurrence in high-risk PFO, whereas there was relative linkage between RLS and stroke recurrence, and no specific factors were associated with stroke recurrence in negative RLS.

There has been considerable interest in risk stratification for stroke recurrence in patients with PFO. With the RoPE (Risk of Paradoxical Embolism) score, higher points, indicating PFO-attributable strokes, predicted a lower stroke recurrence rate.^14^ Meanwhile, the presence of a concomitant ASA increased the risk of stroke recurrence in patients with PFO. Mas et al. analyzed the stroke recurrence rate over four years in 581 patients with cryptogenic stroke ≤55 years of age and with treatment of aspirin 300 mg/day; the stroke recurrence rate was 15.2% in patients with both PFO and ASA, 2.3% in patients with PFO alone, and 4.2% in patients with neither PFO nor ASA.^4^ The present registry study showed that cryptogenic stroke patients with high-risk PFOs had a 1.8 times higher stroke recurrence rate than cryptogenic stroke patients with RLS, but lower than that in the negative RLS group. Compared to the study by Mas et al., the patients enrolled in the present study were older and frequently had atherosclerotic vascular risk factors, leading to the presence of other embologenic diseases such as aortic complicated plaques and non-stenotic carotid plaques, which could be a cause of the higher rate of stroke recurrence in the negative RLS group. More importantly, high-risk PFO could be a risk factor for stroke recurrence in cryptogenic stroke over the entire age spectrum, particularly when comparing with RLS other than with high-risk PFO.

PVH and DSWMH have been shown as degrees of cerebral white matter lesions.^11^ Aging and hypertension are fundamental risk factors for the development of white matter diseases.^11^ Conversely, we previously reported the association of PVH with PFO, and in particular, the co-existence of PFO and ASA was associated with a higher degree of white matter lesions than lone PFO, lone ASA, and neither of them.^15^ Several studies showed that PFO contributed to the development of white matter lesions in Alzheimer’s disease and decompression disease.^16, 17^ Conversely, no such relationship was found in patients with migraine and cerebral autosomal dominant arteriopathy with subcortical infarcts and leukoencephalopathy.^18, 19^ In addition, cerebral perforating arteries can be occluded by small emboli,^20^ even paradoxical emboli.^21^ Thus, small emboli derived from a septal abnormality or the venous system might be related to recurrent stroke and the development of white matter lesions.

Regarding treatment, patients who developed stroke by 2017, when percutaneous PFO closure was not approved in Japan, were enrolled. The frequency of antiplatelet therapy for secondary prevention was higher in the order of negative RLS, RLS, and high-risk PFO groups, whereas the frequency of anticoagulant therapy was the opposite. It is well known that large-scale clinical trials failed to show the efficacy of direct oral anticoagulant (DOAC) therapy for embolic stroke of undetermined source (ESUS),^22, 23^ and a recent systematic review demonstrated that the majority of patients received antiplatelet therapy in ESUS.^24^ There are two possible explanations for selecting anticoagulant therapy in cryptogenic stroke. However, the present registry did not include the presence of deep venous thrombosis, which existed in some patients in the RLS and high-risk PFO groups. Alternatively, some studies indicated that anticoagulant therapy was partially effective for cryptogenic stroke patients.^25, 26^

Some potential limitations of the current study must be considered when interpreting its results. First, due to the retrospective nature of the study design and data extraction procedure, small amounts of data were missing or unavailable. Second, selection bias for the performance of TEE might have been involved in each hospital. Although the protocols for TEE and the interpretations of these findings were common in the two institutions, there was a lack of standardization.

In conclusion, PVH can be a prognostic marker for stroke recurrence in cryptogenic stroke with high-risk PFO, for whom careful monitoring and PFO closure might be considered at <60 years of age.

## Acknowledgments

None

## Data availability statement

The datasets used and analyzed during the current study are available from the corresponding author on reasonable request.

